# Use of Metabolomic Profiling to Understand Variability in Adiposity Changes Following an Intentional Weight Loss Intervention in Older Adults

**DOI:** 10.1101/2020.08.28.20183368

**Authors:** Ellen E. Quillen, Daniel P. Beavers, Anderson O’Brien Cox, Cristina M. Furdui, Jingyun Lee, Ryan M. Miller, Hanzhi Wu, Kristen M. Beavers

**Author notes:** **Corresponding author:** Kristen M. Beavers, Wake Forest University, Winston-Salem, NC 27106.

## Abstract

**Introduction:** Inter-individual response to dietary interventions remains a major challenge to successful weight loss among older adults. This study applied metabolomics technology to identify small molecule signatures associated with loss of fat mass and overall weight in a cohort of older adults on a nutritionally complete, high protein diet.

**Methods:** 102 unique metabolites were measured using LC-MS for 38 adults aged 65-80 years randomized to dietary intervention and 36 controls. Metabolite values were analyzed in both baseline plasma samples and samples collected following the six-month dietary intervention to consider both metabolites that could predict response to diet and those that changed in response to diet or weight loss.

**Results:** Eight metabolites changed over intervention at a nominally-significant level: D-pantothenic acid, L-methionine, nicotinate, aniline, melatonin, deoxycarnitine, 6-deoxy-L-galactose, and 10-hydroxydecanoate. Within the intervention group, there was broad variation in achieved weight-loss and DXA-defined changes in total fat and visceral adipose tissue (VAT) mass. Change in VAT mass was significantly associated with baseline abundance of α-aminoadipate (*p* = 0.0007) and an additional mass spectrometry peak that may represent D-fructose, myo-inositol, mannose, α-D-glucose, allose, D-galactose, D-tagatose, or L-sorbose (*p* = 0.0001).

**Discussion:** This hypothesis-generating study reflects the potential of metabolomic biomarkers for the development of personalized dietary interventions.

## INTRODUCTION

The prevalence of obesity and its detrimental health effects are increasing rapidly among older adults.^1,2^ Medical complications associated with excess fat mass highlight the need to treat obesity in this age group;^3^ yet, recommendation for intentional weight loss remains controversial.^4,5^ Reluctance stems, at least in part, from loss of lean mass known to accompany overall weight loss (10-50% of total tissue^6,7^), and potential exacerbation of age-related disability risk. Encouragingly, data from randomized controlled trials (RCTs) support immediate muscle strength and function gains among older adults following lifestyle-based weight loss – particularly when structured exercise is included as an intervention component – despite lean mass loss.^8,9^

Accordingly, current geriatric obesity treatment guidelines encourage weight loss therapies that minimize lean, while maximizing fat, mass loss for older adults with obesity.^10^ Change in body composition with caloric restriction-induced weight loss appears modifiable through diet, with the amount of dietary protein consumed during caloric restriction identified as a key determinant in lean mass preservation.^11^ Indeed, meta-analytic data show older adults who consume higher levels of protein during weight loss retain more lean mass in comparison with normal protein diets (losses of 21%–22% vs ≥30%).^12^ In agreement, data from our group show older adults following a hypocaloric, nutritionally complete, higher protein meal plan experience smaller average lean (~13%) versus fat (~87%) mass losses; however, the amount and location of fat mass loss was noted to be more variable.^13,14^ Better understanding of the variability in adiposity changes as a means to optimize fat mass loss in this context confers high clinical utility.

Metabolites (i.e., small molecules including sugars, amino acids, and vitamins which are both reactants and products of metabolic processes in the body) are particularly attractive biomarkers for understanding variability in response to dietary interventions. Metabolites can be collected from blood, urine, or other biofluid and are sensitive reflections of both intrinsic and extrinsic changes in nutrition and metabolism.^15,16^ Prior studies have shown that baseline metabolomic profiles are associated with body composition responsiveness^17,18^ and that select metabolites, including branch chain amino acids, may change following a weight loss intervention.^15^ Herein we sought to identify plasma metabolites associated with changes in total and visceral fat mass among older adults following a hypocaloric, nutritionally complete, higher protein meal plan. Although primarily meant to be hypothesis generating, these results may serve as preliminary data to design targeted dietary interventions for fat loss among older adults.

## METHODS

### Study and Participant Descriptions

This analysis utilizes data from The Medifast® for Seniors Study (NCT02730988), a six month RCT conducted at Wake Forest University (recruiting from September 18, 2015 to September 14, 2016), designed to compare the effects of weight loss (WL), achieved by following a hypocaloric, nutritionally complete, higher protein meal plan, versus weight stability (WS) on mobility and body composition in 96 older adults (54-79 years) with obesity (30-40 kg/m^2^). Primary outcome papers, including study design and dietary details, were previously published.^13,14,19,20^ For the present analyses, we evaluated metabolomic data from plasma samples drawn at baseline and at the end of the six-month dietary intervention for 74 individuals with complete DXA-derived body composition data.

### Metabolite Extraction

Metabolites were extracted from 50 μL of plasma spiked with 10 μL of internal standard, 2-(N-morpholino) ethanesulfonic acid (MES) solution using a standard extraction method of four volumes of cold methanol and incubation on ice for 30 minutes. After centrifugation at 18,000 x g for five minutes, the supernatant was dried under vacuum and reconstituted in ultrapure water for liquid chromatography-mass spectrometry (LC-MS) analysis.

### Broad Metabolomics Data Generation and Processing

The LC-MS consisted of a Q Exactive HF hybrid quadrupole-Orbitrap mass spectrometer (Thermo Scientific) and a Vanquish UHPLC system (Thermo Scientific). To identify a broader range of metabolites, samples were analyzed on two different columns, a Hypersil GOLD pentafluorophenyl (PFP) column and an Accucore Vanquish C18+ column. A linear gradient was employed for chromatographic separation using 100% water (mobile phase A) and 90% acetonitrile (mobile phase B) both of which contained 0.1% formic acid and 10mM ammonium formate. Data was acquired by collecting full mass spectra (MS1) using polarity switching (positive/negative) at a resolution of 150K. Unnormalized metabolite data files are available at NIH Common Fund’s National Metabolomics Data Repository https://www.metabolomicsworkbench.org).

MS peaks were extracted and integrated within the MSMLS Discovery software (IROA Technologies) in combination with customized compound libraries prepared using a Mass Spectrometry Metabolite Library (Sigma-Aldrich). To eliminate redundancy in compound identification, the most abundant ion in each peak was selected and peak area was then normalized to the total ion current (TIC) for relative quantification. We quantile normalized and log transformed the data to reduce the bias caused by a small number of highly abundant metabolites.^21^ Because missing data from a mass spectrometer is not generally representative of a true zero where values are present for other samples, we replaced missing values with half of the minimum detected value in the full data set.

### Body Weight, Composition, and Fat-Distribution

Body weight and composition were measured at baseline and six months. Baseline weight was measured to the 10th decimal without shoes and outer garments using a calibrated scale (Detecto 758C Weight Indicator; Webb City, MO). Height was obtained without shoes to within 0.25 inches using a QuickMedical 235D Heightronic Digital Stadiometer (Issaquah, WA). Dual-energy x-ray absorptiometry (DXA) scans were used to determine total body, lean, and fat masses, with visceral adipose tissue (VAT) derived using the CoreScan algorithm (GE Medical Systems, Madison, WI, USA).^22^ All scans were performed on the same machine and by the same technician, following manufacturer recommendations for patient preparation and positioning. Coefficients of variation from repeated measurements at our institution are <1.0% for total body, lean, and fat masses.

### Statistical Analyses

For participants with complete DXA data (n=74), baseline metabolomic data was evaluated using principal components analysis to identify stratification associated with randomization assignment, age, gender, race, or technical artifacts (i.e., LC-MS run order). Demographic characteristics, including age, gender, and race were assessed by participant report at baseline.

For 102 unique MS peaks, mean differences in the WL and WS groups were evaluated using Welch’s t-tests to account for differences in variance. Within the WL group (n=38), we evaluated differences in loss of total weight, total fat mass, and VAT mass associated with either metabolite abundance at baseline or change in metabolite abundance over the six-month intervention using linear regression.

We also considered percent change in weight and body composition to account for baseline differences. Change in weight, total fat mass, and VAT mass all showed substantial but non-significant differences between men and women at baseline, so we considered both models containing all participants and gender-stratified models. Males were 23.5 kg heavier on average with 3.6 kg more total fat mass and 1.9 kg more VAT mass. The relationship of demographic variables with individual metabolites and principal components of metabolomic variation were tested and found to be non-significant except with regard to gender. Based on this, age and race were not incorporated into subsequent models. We undertook these analyses solely in the WL group because the diet provided to this group was expected to cause broad shifts in the nutrient profile irrespective of weight loss that could confound associations if the WS group was included. All analyses were performed in R (R Foundation for Statistical Computing, Vienna, Austria).^23^ Throughout, MS peaks were considered independent of one another so multiple testing is addressed through the use of Bonferroni corrected critical p-values.

## RESULTS

### Intervention Related Changes in Body Composition and Sample Characteristics

Demographic and body composition data for the subset of individuals with complete metabolomics and body composition data are presented in **Table 1**. Overall, participants were 65-80 years of age, 58% female, and 76% white with starting BMI between 30 and 42 42 kg/m^2^. Intervention related change in total and regional body composition were published previously.^13,14^ Briefly, over the course of the six month period, total body mass was significantly reduced in the WL group [-8.2 kg (−9.6, −6.8)] as compared with the WS group [-1.2 kg (−2.6, 0.3)], with 87% of total mass lost as fat [WL: −7.1 kg (−8.1, −6.1) kg; −16% change from baseline]. Significant reductions in VAT were also observed in the WL [-1.9 kg (−1.7, −2.1); – 20.1%], but not WS [2.3 kg (2.2, 2.5); 2.1%], group.^14^ In contrast, lean mass loss was much more modest, and no differential treatment effect was observed between groups [WL: −0.8 kg (−1.4, – 0.2] vs WS: −0.2 kg (−0.9, 0.4) kg]. **Figure 1** highlights the broad distribution in weight, total fat, and VAT mass lost over the intervention compared to change in lean mass in the current study sample (N = 74).

**Table 1.**
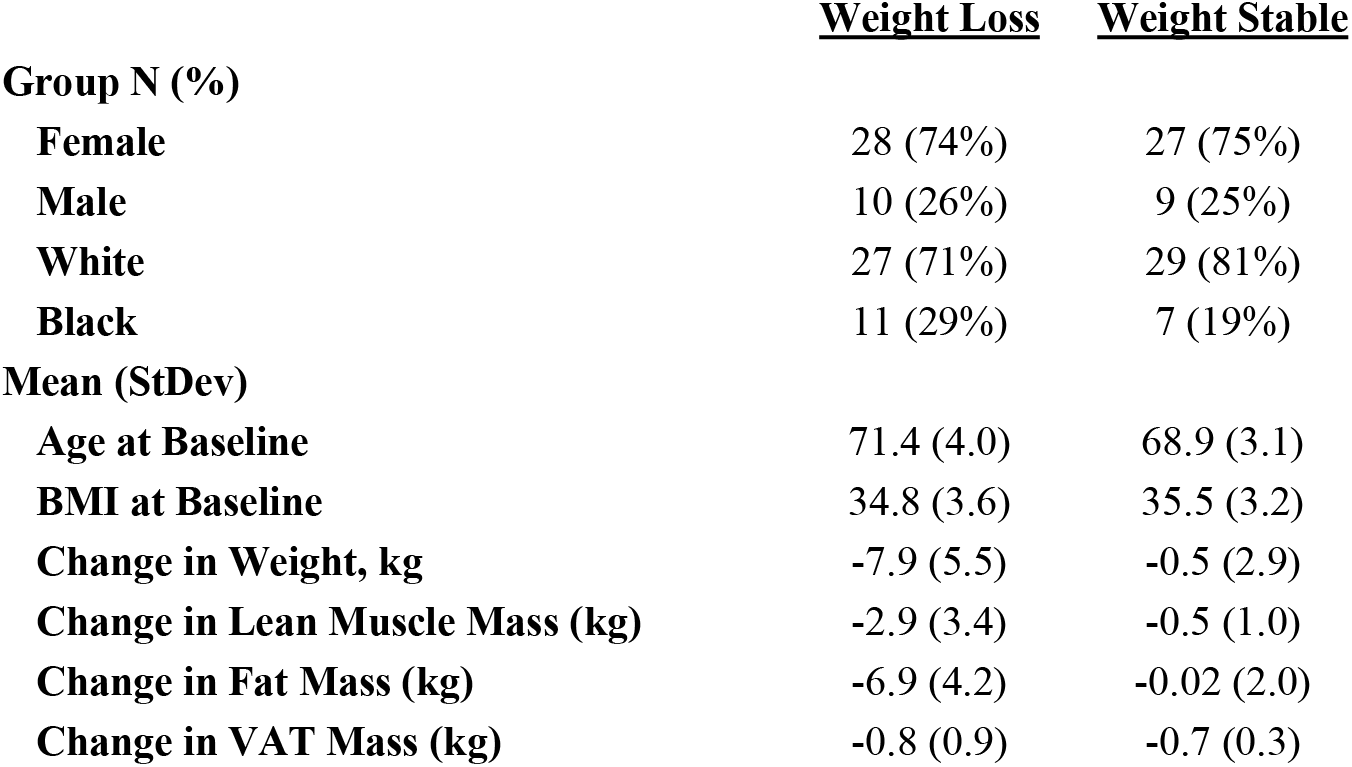
Demographic characteristics and baseline and six-month change in body composition for weight loss and weight stable groups.

**Figure 1.**
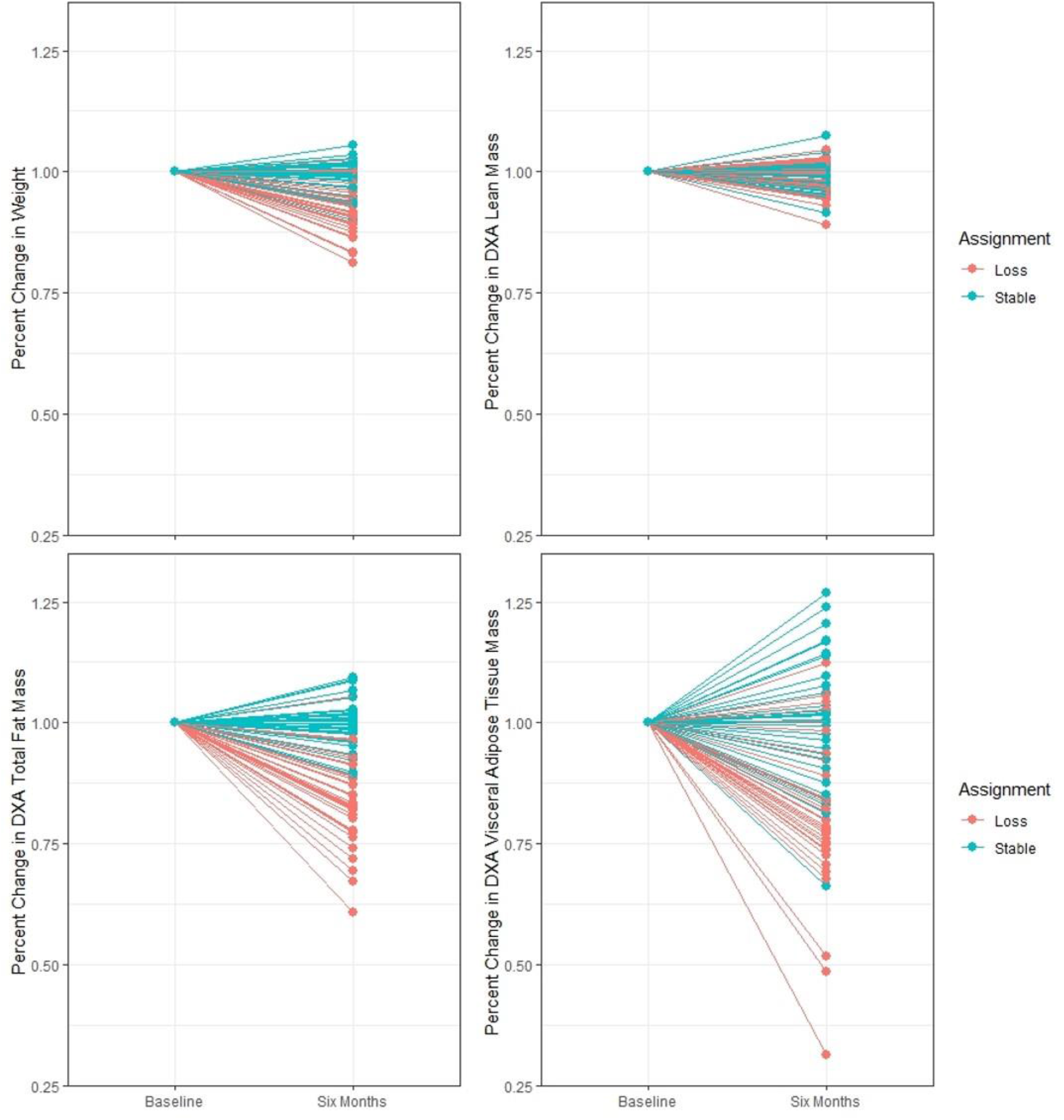
Change in Weight and Body Composition. From baseline to six months, the percent change in total body weight, lean mass, fat mass, and VAT mass in individuals randomized to the weight loss (orange; n=38) and weight stable (teal; n=36) groups.

### Metabolomics Data Generation and Normalization

We detected 102 unique mass spectrometry features (MS1 peaks) peaks present above background levels in at least 10 samples corresponding to 118 metabolites due to some overlap in chromatographic separation. After normalization, three individuals were removed from the sample for non-standard distributions of metabolites identified in the principal components analysis (PCA). This appears to have been driven by the number of missing metabolites in these samples.

### Metabolites Associated with Participant Baseline Characteristics

In the baseline plasma samples, we identified only two metabolites (creatine and 5,6-dihydrouracil) differing by gender, and none that differ by age or race. We note that all participants are between 65 and 80 years old, so the effect of age on metabolite variability will be muted.

### Metabolomic Differences in Weight Loss vs. Weight Stable

No significant differences in metabolite abundance were found between the WL and WS groups at baseline. After six months on the intervention diet, five metabolites – D-pantothenic acid, L-methionine, nicotinate, aniline, and melatonin – decreased in the WL group but not the WS group while three metabolites increased – deoxycarnitine, 6-deoxy-L-galactose, and 10-hydroxydecanoate using a nominal α = 0.05 (**Figure 2**). The heatmap in **Figure 3** shows the clustering of WL and WS plasma samples and metabolites at six months based on differentially expressed metabolites. We note that the clustering of the WL and WS groups are not mutually exclusive across the top axis.

**Figure 2.**
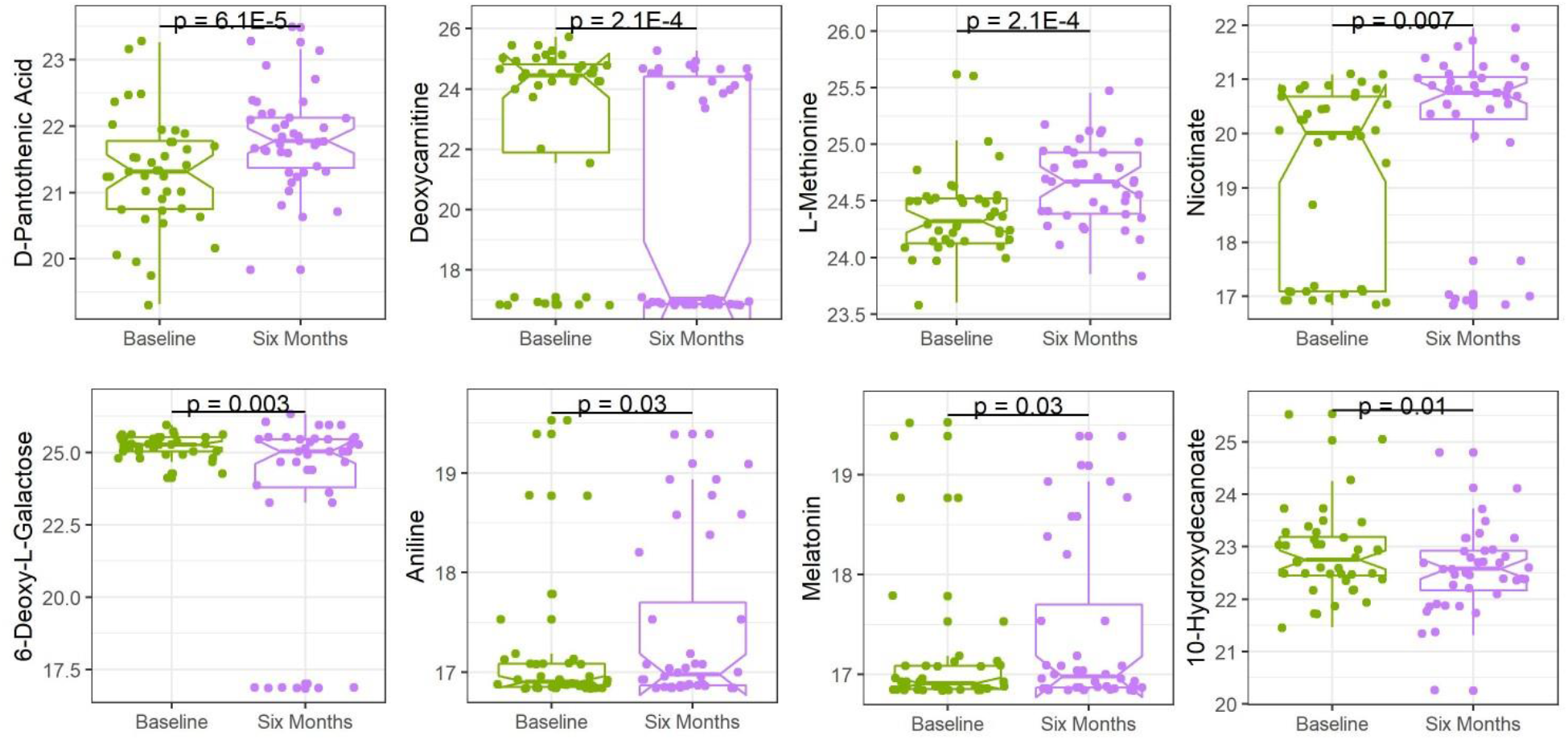
Change in Mean Metabolite Abundance in Weight Loss Group. The distribution with mean and inter-quartile ranges are shown for eight metabolites with nominally significant p-values in Welch’s t-tests between baseline and the end of the six-month weight loss intervention.

**Figure 3.**
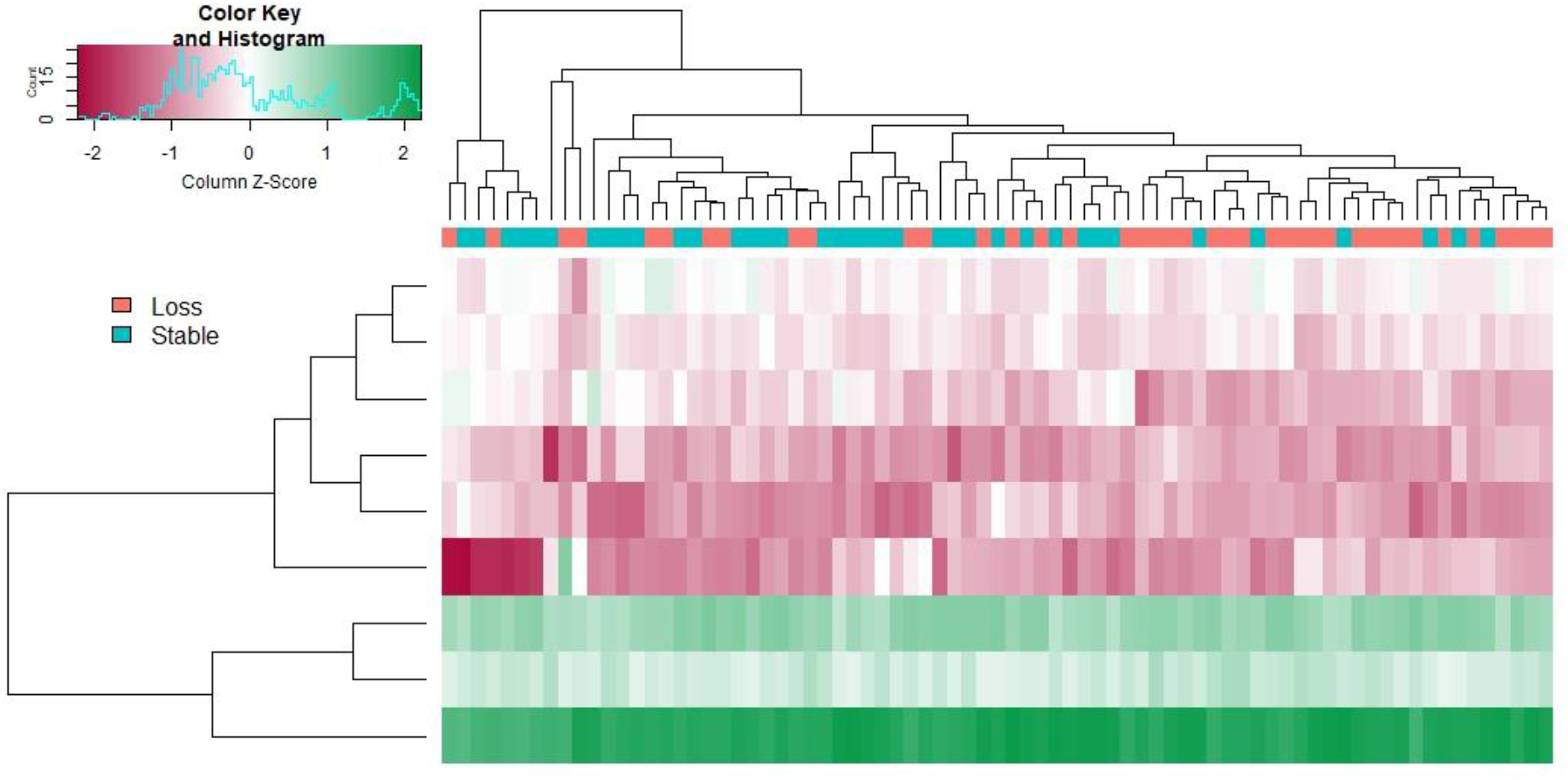
Heatmap of Differentially Expressed Metabolites between Weight Loss and Weight Stable Groups at End of Intervention. Based on metabolites with nominally significant (*p* < 0.05) Welch’s t-tests comparing WL and WS groups at the six-month time point, this heatmap shows clustering of highly expressed (green) and lowly expressed (red) metabolites. The dendrogram at the top clusters individuals based on the similarity of their metabolite profiles with individuals in the WL group in orange and WS group in teal

### Metabolites Associated with Change in Weight and Fat Mass

We evaluated change in metabolite abundance for all 102 MS1 peaks from baseline to six months versus percent change in weight, total fat mass, or VAT to evaluate metabolites that may be produced during weight loss. At a Bonferroni-correct *p* = 0.0007, no metabolites were significantly associated.

When comparing percent change in weight and fat mass with baseline variables to identify potential predictive biomarkers, one MS peak was marginally significant in the full analysis, α-aminoadipate (R^2^ = 0.29, *p* = 0.0007). This peak was also nominally significant in the female-specific analysis. In the female-specific analysis, metabolite abundance at one MS peak is significantly associated at *p* = 0.0001 (R^2^ = 0.46). This peak is linked to eight potential metabolites present in the Mass Spectrometry Metabolite Library (D-fructose, myo-inositol, mannose, α-D-glucose, allose, D-galactose, D-tagatose, or L-sorbose), which cannot be distinguished by accurate mass or chromatography profiles. **Figure 4** shows the relationship between each peak intensity in WL participants at baseline versus VAT change over intervention.

**Figure 4.**
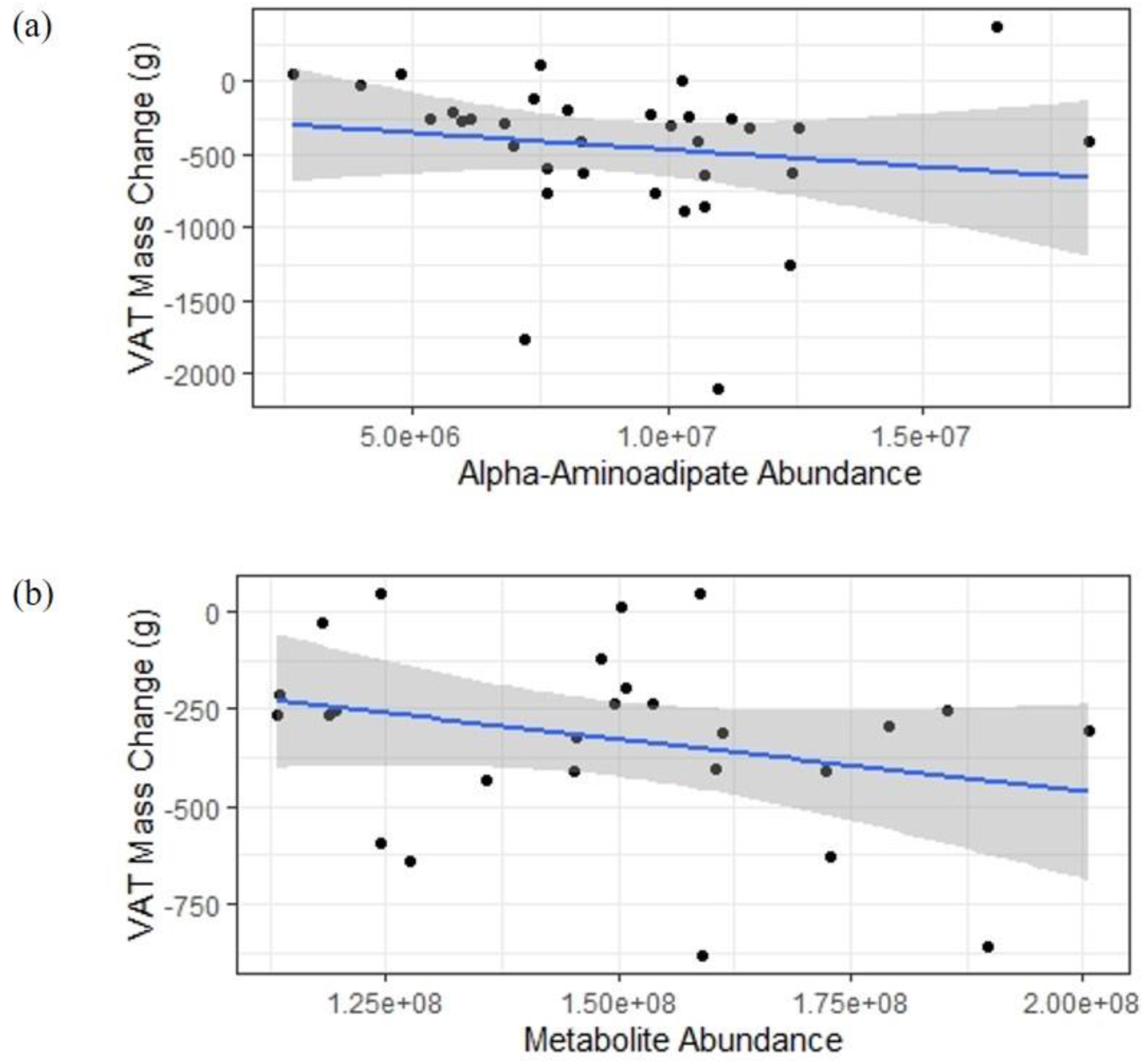
Scatter Plot of Baseline Metabolite Abundance vs. Change in VAT. (a) Abundance of α-aminoadipate at baseline in all individuals randomized to WL vs. VAT mass change in grams following intervention. (b) Abundance of the indeterminate metabolite at baseline vs. VAT mass change for females in the WL group only.

## DISCUSSION

The goal of this study was to identify metabolites associated with inter-individual differences in total and regional fat loss during a weight loss intervention in older adults known to produce a lean mass sparing effect. When compared to the weight stable group, three metabolites increased across the intervention (deoxycarnitine, 6-deoxy-l-galactose, and 10-hydroxydecanoate) while five metabolites decreased (D-pantothenic acid, L-methionine,nicotinate, aniline, and melatonin). Additionally, we report an association between greater baseline levels of α-aminoadipate (also called 2-aminoadipic acid) and myo-inositol or carbohydrate metabolites (α-D-glucose, allose, mannose, D-galactose, D-fructose, and L-sorbose) with larger decreases in visceral adipose tissue. This information is a critical first step in developing personalized approaches to optimize weight loss interventions for older adults.

Given the dearth of literature concerning the metabolites that change in response to change in diet, it is difficult to hypothesize about the origins of diet-associated metabolite increases that were observed in our study, but is possible they are related to the long-term change in diet (e.g., caloric restriction or dietary composition). Interestingly, deoxycarnitine has previously been shown to increase following weight loss interventions in animal models.^24^ Since deoxycarnitine is the final intermediate for endogenous L-carnitine biosynthesis, it is possible that L-carnitine biosynthesis was elevated in the weight loss group, resulting in greater decreases in visceral adipose tissue.^25^

Observed decreases in amino acids support previous observations and may reflect improvements in metabolic perturbations.^26-28^ However, despite observing decreases in amino acids and visceral adipose tissue in the weight loss group, previous research examining the relationship between visceral fat depots and amino acids remains unclear and were not identified in our study.^29-31^ The weight loss group also displayed declines in vitamin B family metabolites (pantothenic acid and nicotinate), which may have been a function of decreased food intake as part of enrolling in the intervention. Interestingly, decreases in melatonin occurred within the weight loss group, which has displayed promising effects for improving body composition and may ameliorate the consequences of obesity.^32-34^

To date, few studies have examined if baseline metabolomic signatures are associated with weight loss outcomes.^17,18^ Stroeve et al.^17^ suggested that 57% of weight loss success could be identified from baseline metabolomic signatures in morbidly obese men and women, which included acetoacetate, triacylglycerols, phosphatidylcholines, amino acids, creatine and creatinine. Additional observations suggest that baseline xylitol and uridine levels were inversely associated with weight loss, whereas 2-aminobutyric acid and glyceric acid were positively associated with weight loss ≥10% over a 1-year intervention.^18^

The metabolites identified in our study mirror and extend recent observations between these metabolites (e.g., myo-inositol and glycolytic/gluconeogenic intermediates) and visceral adipose tissue.^35^ When previous research stratified weight loss participants into high-or low-responders post-intervention (≥7.2 kg and ≤5.2 kg weight loss, respectively); myo-inositol, among other metabolite levels, were greater in the high-responders at baseline.^36^ Additionally, previous research examining myo-inositol supplementation has also observed an attenuation of metabolic abnormalities.^37-39^ Regarding the elevated carbohydrate metabolites, obesity has been shown to be associated with increased levels of these compounds, and the metabolomic signature of carbohydrate metabolism for obesity and diabetes are quite similar.^40^ This is further bolstered by our association of elevated α-aminoadipate levels at baseline with greater VAT mass loss. This metabolite is an oxidation product considered a biomarker of early stage diabetes and for cardiometabolic complications among individuals with diabetes.^41,42^

Personalized interventions to maximize overall improvements in health is the ultimate goal of weight-loss research in older adults. Because of its association with increased risk of cardiometabolic diseases^43^ (independent of BMI), identification of biomarkers associated with loss of VAT may be particularly important in targeting dietary approaches that can minimize lean mass loss while maximizing cardiometabolic benefit. Given the previous links between visceral adipose tissue and altered metabolic effects,^35,44^ the elevated carbohydrate metabolites at baseline may identify individuals with greater metabolic irregularity (e.g., those with decreased metabolic health). While no study participants had diabetes, these baseline metabolites may represent individuals with the most to gain cardiometabolically from a weight loss intervention.

While studies of weight-loss associated metabolomic biomarkers have increased rapidly in recent years, we are among a limited number of studies to report baseline metabolite profiles associated with response to dietary interventions.^15,17,18^ Our findings are limited by the fact that analyses were performed post hoc and the study was not originally designed or powered to evaluate metabolite associations. However, the identification of baseline glucose metabolites linked to variation in VAT loss provides an avenue to identify individuals who may benefit most from a lower-sugar dietary prescription. Future studies incorporating lipidomics or untargeted metabolomics could further elucidate differences at baseline that predict response to interventions.

## Data Availability

Submission of metabolomic data to the NIH metabolomics workbench is pending.

## ACKNOWLEDGMENTS

This work was supported by a grant from Jason Pharmaceuticals, Inc., a wholly owned subsidiary of Medifast, Inc., as well as the Wake Forest Claude D. Pepper Older Americans Independence Center (P30 AG21332) and Proteomics and Metabolomics Shared Resource Core (P30CA012197). Support was also provided by the National Institute on Aging through an exploratory and developmental grant (R21 AG061344) to KMB, DPB, and CF, career development award (K01 AG056663) to EQ and postdoctoral fellowship (T32 AG033534) to RM. The content is solely the responsibility of the authors and does not necessarily represent the official views of the National Institute of Health.

## AUTHOR CONTRIBUTIONS

Concept and design (KMB, DPB); acquisition of data (KMB, CF, JL, AOC, HW); analysis and interpretation of data (EQ, JL), drafting of manuscript (KMB, EQ, RM), critical revision (CF, DPB). CF takes responsibility for the accuracy and integrity of mass spectrometry data. EQ had full access to all the data in the study and takes responsibility for the accuracy of the data analysis. All authors approve the final version of the manuscript.

## CONFLICTS OF INTEREST

Medifast, Inc. provided partial funding for the study and made an in-kind product donation for the meal replacements used in the study. The terms of this arrangement were reviewed and approved by Wake Forest University Health Sciences in accordance with its conflict of interest policies.

## Notes

### Author Declarations

Wake Forest School of Medicine IRB

